# Age at Menarche and Coronary Artery Disease Risk: Divergent Associations with Different Sources of Variation

**DOI:** 10.1101/2024.08.14.24312022

**Authors:** Ambreen Sonawalla, Daniel I. Chasman, Yee-Ming Chan

## Abstract

**Background:** In women, both earlier and later age at menarche (AAM) are associated with increased risk of coronary artery disease (CAD). This study sought to determine if the relationship of AAM with CAD and CAD risk factors differs for different underlying sources of variation in AAM – specifically, variation attributable to common genetic variants as represented by a polygenic score (PGS) vs. variation in AAM adjusted for the PGS.

**Methods:** Primary analyses were conducted on data from 201,037 women in the UK Biobank and validation studies on data from 23,268 women in the Women’s Genome Health Study (WGHS). For each individual, a PGS for AAM was calculated, then two variables were estimated from linear regression models: the genetically predicted AAM (the estimated AAM for each woman solely due to the effects of common genetic variants) and the PGS-adjusted AAM (estimated AAM for each woman solely due to factors other than the PGS). Logistic regression and linear splines were then used to study the relationships of these variables with CAD and CAD risk factors.

**Results:** Genetically predicted AAM demonstrated a linear relationship with CAD and linear or roughly linear relationships with CAD risk factors. In contrast, PGS-adjusted AAM demonstrated a U-shaped relationship with CAD and with hemoglobin A1c, triglycerides, HDL-C, and waist-hip ratio. Validation studies using WGHS data produced similar results.

**Conclusions:** These results suggest that later AAM itself does not cause increased risk of CAD; rather, upstream sources of variation other than common genetic variants can cause both later AAM and increased risk of CAD. Dysglycemia, dyslipidemia, and central adiposity are candidate mediators of the association of later AAM with increased risk of CAD.

## INTRODUCTION

Many pathologies manifesting in adulthood have antecedents in childhood. There is growing evidence that coronary artery disease (CAD) in women, a leading cause of morbidity and mortality in the world, is associated with both earlier and later age at menarche (AAM), a hallmark of pubertal timing (1–4). *Earlier* puberty is associated with increased risk of CAD in both men and women; however, the association between *later* puberty and increased risk of CAD appears to be unique to women; in men, later puberty is associated with a decreased risk of CAD (3). A deeper exploration of these childhood antecedents would allow a better understanding of the pathogenesis of CAD in adulthood, specifically identify factors that uniquely affect women and facilitate the development of targeted preventive interventions, potentially as early as childhood.

Multiple studies have associated earlier AAM with a higher risk of developing components of the metabolic syndrome, namely obesity, type 2 diabetes mellitus, hypertension, and dyslipidemia (3,5–9). Studies have further suggested that the association between earlier AAM and risk of CAD is mediated by adiposity (10,11). In contrast, the associations of later AAM reported to date do not fit neatly into the paradigm of metabolic syndrome. Later AAM is associated with lower rather than higher body-mass index (BMI) (3,12,13), and studies on other components of the metabolic syndrome have produced conflicting results, with some studies showing association of later AAM with higher risk of hypertension (1,14), others showing a lower risk of hypertension (3) or type 2 diabetes (15), and yet others showing no association with hypertension, type 2 diabetes, or hypercholesterolemia (3,16). Hence, while earlier AAM has been associated with several CAD risk factors, there may be distinct mechanisms underlying the association of later AAM with increased risk of CAD.

Variation in AAM can stem from several upstream sources, including genetics (both common genetic variants and rare genetic variants), acquired factors such as chronic illness, chronic stress, underweight, and undernutrition, and environmental factors such as family composition (e.g., presence or absence of father) (17). It is possible that some of these sources of variation may influence risk of CAD only through their influence on AAM, while others may directly influence CAD risk and features of the metabolic syndrome (Figure 1). Thus, dissecting variation in AAM based on underlying sources of variation could provide a clearer understanding of the relationship of AAM with CAD.

**Figure 1:**
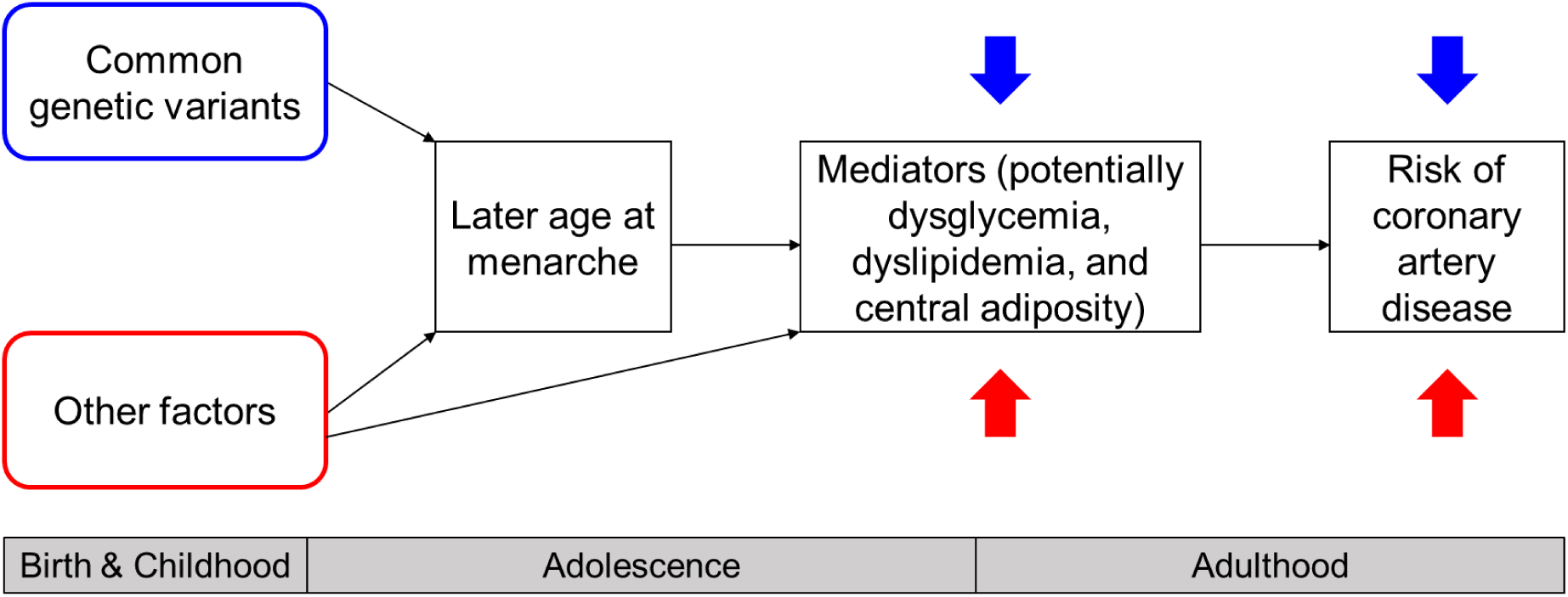
Relationships of different sources of variation in age at menarche (AAM) with coronary artery disease (CAD) and CAD risk factors. Variation in AAM was dissected into variation associated with common genetic variants (as estimated by a polygenic score for AAM) and variation independent of the polygenic score, then associations of these sources of variation with CAD and CAD risk factors were studied. When associated with common genetic variants, later AAM was associated with a lower risk of CAD and favorable changes in CAD risk factors (blue arrows). However, when occurring independently of the polygenic score, later AAM was associated with higher risk of CAD and unfavorable changes in CAD risk factors (red arrows). The “other factors” causing later AAM independent of the PGS have yet to be identified and could potentially include chronic illness, underweight, and rare genetic variants.

Genetics is a major source of variation in AAM, with half to three-quarters of variation attributable to genetics (18,19). A 2017 genome-wide association study (GWAS) on AAM identified 389 independent single-nucleotide polymorphisms (SNPs) associated with AAM at genome-wide significance (20). The results from this GWAS allow the calculation of a polygenic score (PGS) for a given individual to reflect the cumulative contribution of common genetic variants to AAM.

Previous studies have used PGSs to dissect the influence of genetics vs. environmental factors (or other factors not captured by the PGS) that contribute to traits such as BMI and LDL-C (21,22). These studies have found that associations of these traits with health outcomes differ between genetically and environmentally influenced traits. For example, obesity driven by environmental factors was associated with more harmful cardiovascular outcomes than genetically predicted obesity (21), suggesting that dissecting effects based on underlying source of variation can allow deeper insights into pathogenic mechanisms.

The aims of this study were two-fold: first, to determine if the association between later AAM and increased risk of CAD depends on the underlying source of variation in AAM – specifically, common genetic variation vs. other sources of variation; second, to study the relationships of these different sources of variation in AAM with CAD risk factors.

## METHODS

To study how different sources of variation in the timing of menarche are associated with risk of CAD, two variables were calculated: genetically predicted AAM (the estimated AAM determined solely by the effects of common genetic influences, as estimated by a PGS for AAM) and PGS-adjusted AAM (the estimated AAM determined solely by factors other than common genetic variants, the PGS).

### Study cohorts

This study used data from two cohorts: the UK Biobank for primary analyses and the Women’s Genome Health Study (WGHS) for validation analyses. The UK Biobank is a population-based cohort of over 500,000 men and women in the UK 40 years and older at the time of recruitment, with extensive health-related phenotypic and laboratory data as well as individual-level genetic data (23). This study analyzed data from 201,037 unrelated women in the UK Biobank of non-Finnish European ancestry (as determined through principal component analysis) (24) who had genetic and self-reported AAM data. During data collection in the UK Biobank, any value of AAM <5 years or >25 years was rejected, and any AAM entered as <6 years or >20 years required confirmation from participants (25). Women with missing AAM data were excluded from analysis. The UK Biobank obtained the multiple ethical and regulatory approvals required for recruitment and research procedures, and participants provided written consent (23). The WGHS is a cohort of initially healthy American women aged 45 years and older at enrollment with genetic and phenotypic data, followed over 26-28 years for cardiovascular and other outcomes (26). This analysis studied data from 23,268 women in the WGHS who had genetic and self-reported AAM data. Self-reported AAM ≤9 years or ≥ 17 years were entered as 9 years or 17 years, respectively. The WGHS was approved by the institutional review board of Brigham and Women’s Hospital, and participants consented to ongoing analyses (26).

### Polygenic score calculation

A PGS was calculated for each woman in the above cohorts in two steps. The first step used the PRS-CS algorithm, which allows the inclusion of all available SNPs from the GWAS on AAM (20), not just those that meet a given *p*-value threshold, and weights the SNP effect sizes based on their significance and adjusts for linkage disequilibrium (27). The use of this algorithm has the potential to explain more variability in AAM than algorithms that use only the SNPs that meet a given significance threshold. The second step calculated the PGS using PRSice-2 (without clumping or thresholding) to sum the weighted effect sizes for all SNPs in each individual with the ability to incorporate the probabilistic genotype dosages generated by imputation (28).

### Subdividing variation in age at menarche

Using each full cohort, regression of self-reported AAM was performed against the PGS for AAM, with the first 10 genetic principal components, assessment center and technical variables such as array number as covariates. The following two variables were then calculated for each individual:

1. Genetically predicted AAM: This represents the estimated AAM that each woman would have had if her AAM were determined solely by her common genetic variants, as estimated by the PGS. Statistically, it is the AAM predicted by the regression of AAM against the PGS (Supplemental Figure 1, (29)).
2. PGS-adjusted AAM: This represents the estimated AAM that each woman would have had if there were no effect of her common genetic variants (as represented by the PGS), i.e., her AAM were determined solely by sources of variation other than common genetic variants. Statistically, this was the residual of the regression of AAM against the PGS for each individual, added to the AAM corresponding to the mean PGS (to simulate a scenario in which the contribution of the PGS is the same for all women) (Supplemental Figure 1, (29)).

### Outcomes and analytical methods

The study’s primary outcome variable was CAD risk. In the UK Biobank, prevalent CAD at baseline was determined as previously described using a combination of self-report, ICD-9/10 codes, and procedure codes (24). The WGHS recruited middle-aged female healthcare professionals with no history of CAD at baseline and identified validated incident CAD during 26-28 years of follow-up as described previously (26). Secondary outcome variables were CAD risk factors at baseline for both cohorts: hemoglobin A1c (HbA1c), triglycerides, high-density lipoprotein cholesterol (HDL-C), low-density lipoprotein cholesterol (LDL-C), systolic blood pressure (SBP), diastolic blood pressure (DBP), and BMI; data on waist-hip ratio was available at baseline in the UK Biobank and 6 years after recruitment in the WGHS.

The relationship of genetically predicted AAM and PGS-adjusted AAM was studied with logistic regression for prevalent CAD, with Cox proportional hazards models for incident CAD, and with linear splines for each continuous variable (HbA1c, triglycerides, HDL-C, LDL-C, SBP, DBP, BMI, waist-hip ratio), with a knot at 12.94 years, which is the AAM corresponding to the mean PGS. Because we observed nonlinear relationships, we also used linear splines to separately analyze values of genetically predicted and PGS-adjusted AAM earlier and later than the mean. For analyses with LDL-C, results were corrected for self-reported use of cholesterol-lowering medications – these were specific LDL-lowering medications in the UK Biobank, (statins, ezetimibe, and bile-acid sequestrants) and collective cholesterol-lowering medications in the WGHS; additional analyses included only women not taking these medications.

Covariates of age and age^2^ were used for all analyses. Analyses were conducted using R v.4.3.1. A significance threshold of 0.05 was used.

To determine if results in the UK Biobank were biased by overfitting as a result of the UK Biobank having contributed to the GWAS for AAM, analyses of CAD were repeated using a PGS calculated using an earlier GWAS that did not include the UK Biobank (30). Furthermore, to determine if outlier values of AAM were disproportionately affecting results, analyses were repeated after excluding women with extreme values of AAM such that up to 0.1% of women were excluded at each extreme.

## RESULTS

To understand potential different effects of different sources of variation in AAM, we constructed two hypothetical scenarios. In the first scenario, a woman’s AAM is determined solely by the effects of common genetic variants. We refer to the estimated AAM in this scenario as “genetically predicted AAM.” In the second scenario, a woman’s AAM is determined solely by the effects of factors other than common genetic variants (or more precisely, by factors other than the PGS). We refer to the estimated AAM in this scenario as “PGS-adjusted AAM.”

To examine the differential associations of these two sources of variation with CAD risk, we analyzed data from 201,037 unrelated, non-Finnish European women in the UK Biobank. We first calculated a PGS for AAM for each woman, then regressed self-reported AAM against the PGS for AAM to calculate genetically predicted AAM and to derive PGS-adjusted AAM (Supplemental Figure 1, (29)). For instance, for an individual with a self-reported AAM of 16 years and a PGS of 0.357, the regression provided a genetically predicted AAM of 13.42 years and a residual of 2.58 years. In the regression, the AAM corresponding to the mean PGS was 12.95 years, and hence the PGS-adjusted AAM was 2.58 + 12.95 = 15.53 years.

For women in the UK Biobank, the regression of self-reported AAM against the PGS for AAM demonstrated that the PGS accounted for 15.8% of the variation in AAM. Genetically predicted AAM had a mean ± standard deviation of 12.95 ± 0.64 years, and PGS-adjusted AAM had a mean ± standard deviation of 12.94 ± 1.47 years with a standard deviation of 1.47 years (Supplemental Figure 2, (29)). We then studied the associations of genetically predicted AAM and PGS-adjusted AAM with risk of CAD and with CAD risk factors.

### Risk of coronary artery disease

Risk of CAD demonstrated a linear relationship with genetically predicted AAM but a non-linear, U-shaped relationship with PGS-adjusted AAM (Figure 2). In the linear relationship of genetically predicted AAM with risk of CAD; each 1-year increase in genetically predicted AAM was associated with an odds ratio (OR) for CAD of 0.91. In other words, for every 1 year that AAM was later due solely to the effects of common genetic variants (as estimated by the PGS), the odds of CAD were lower by 9% (Table 1). To assess for non-linear relationships, linear spline analyses were done which showed no difference in slopes when genetically predicted AAM was earlier vs. later than the mean (Table 1). This lack of difference in slopes indicates that the linear relationship of genetically predicted AAM and risk of CAD extends across all values of genetically predicted AAM.

**Figure 2:**
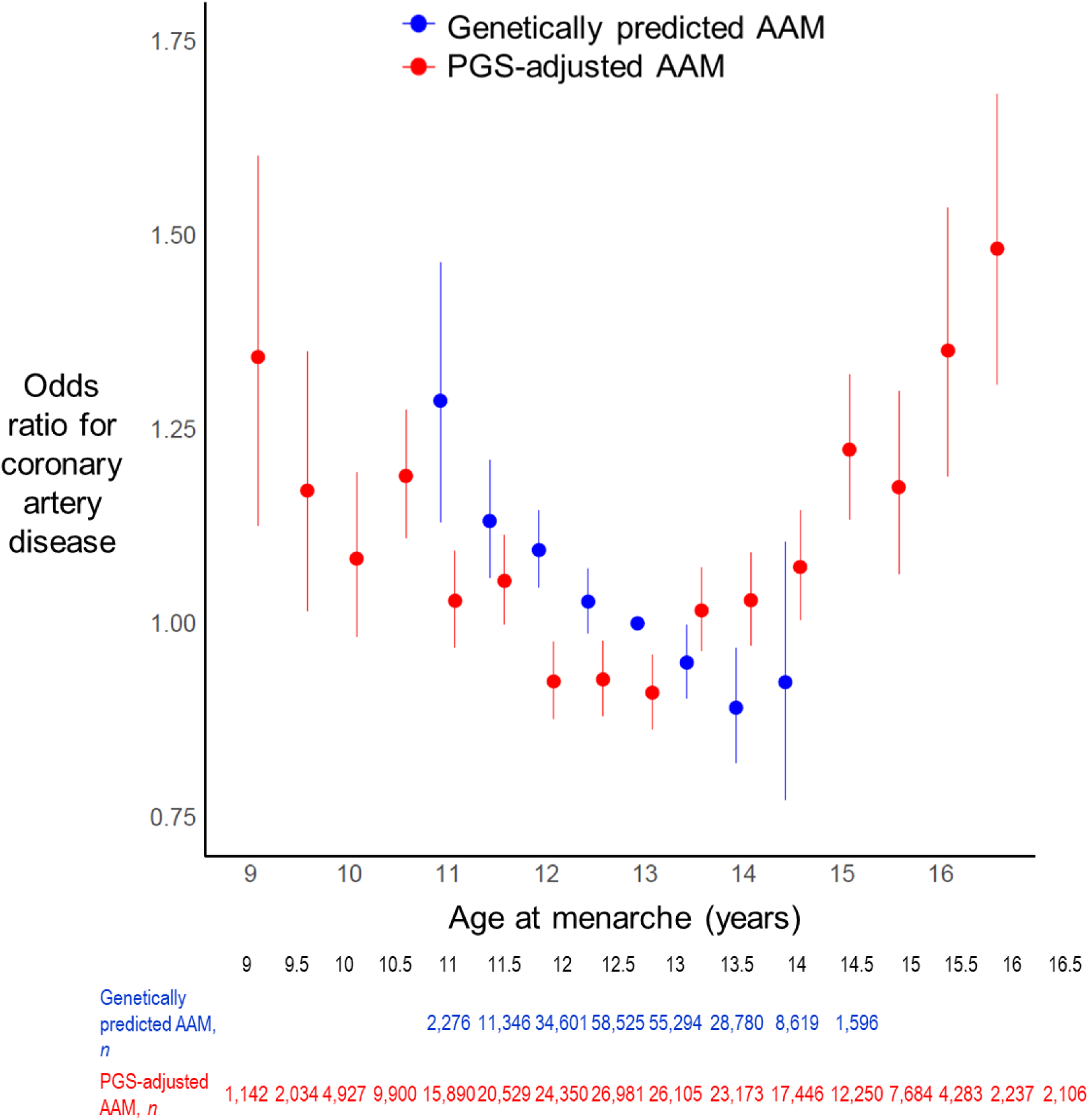
Associations of different sources of variation in age at menarche (AAM) with odds of coronary artery disease (CAD) in the UK Biobank. For each individual in the cohort, variation in AAM was dissected into variation attributable to common genetic variants as estimated by a polygenic score (PGS) for AAM (represented by “genetically predicted AAM”) and variation attributable to sources other than common genetic variants (represented by “PGS-adjusted AAM”), then associations with risk of CAD were studied. Genetically predicted AAM showed a linear association with risk of CAD, whereas PGS-adjusted AAM showed a U-shaped association. The different associations of genetically predicted AAM and PGS-adjusted AAM with risk of CAD suggest that it’s not later AAM itself that causes increased risk of CAD, but that factors other than the PGS can cause both later AAM and increased risk of CAD.. To achieve bin sizes of ≥1000 individuals, the first and last bins for each variable represent the group of individuals with AAM ≤ or ≥ the value listed on the x-axis, respectively. Dots represent estimates; bars represent standard errors.

**Table 1:**
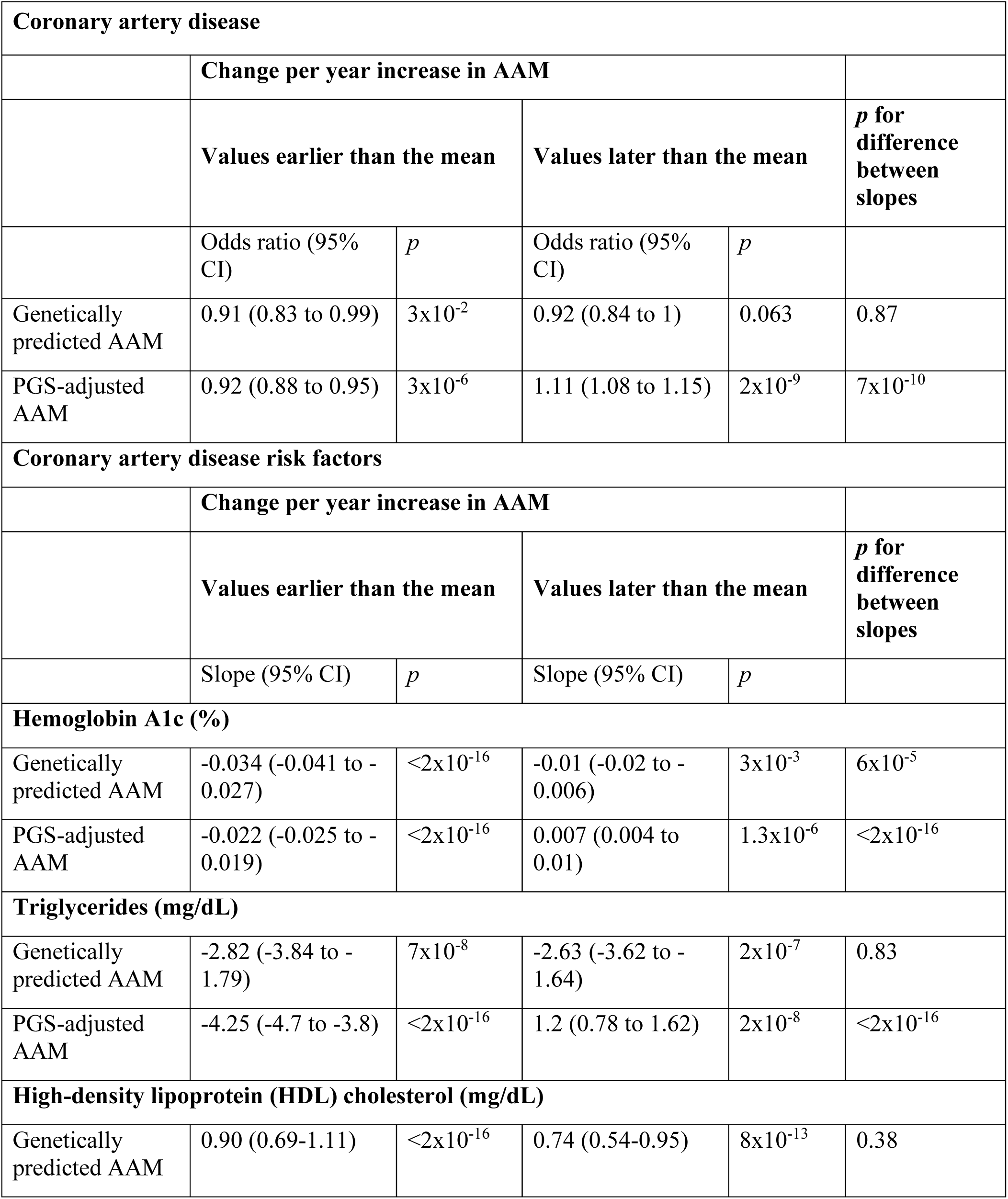
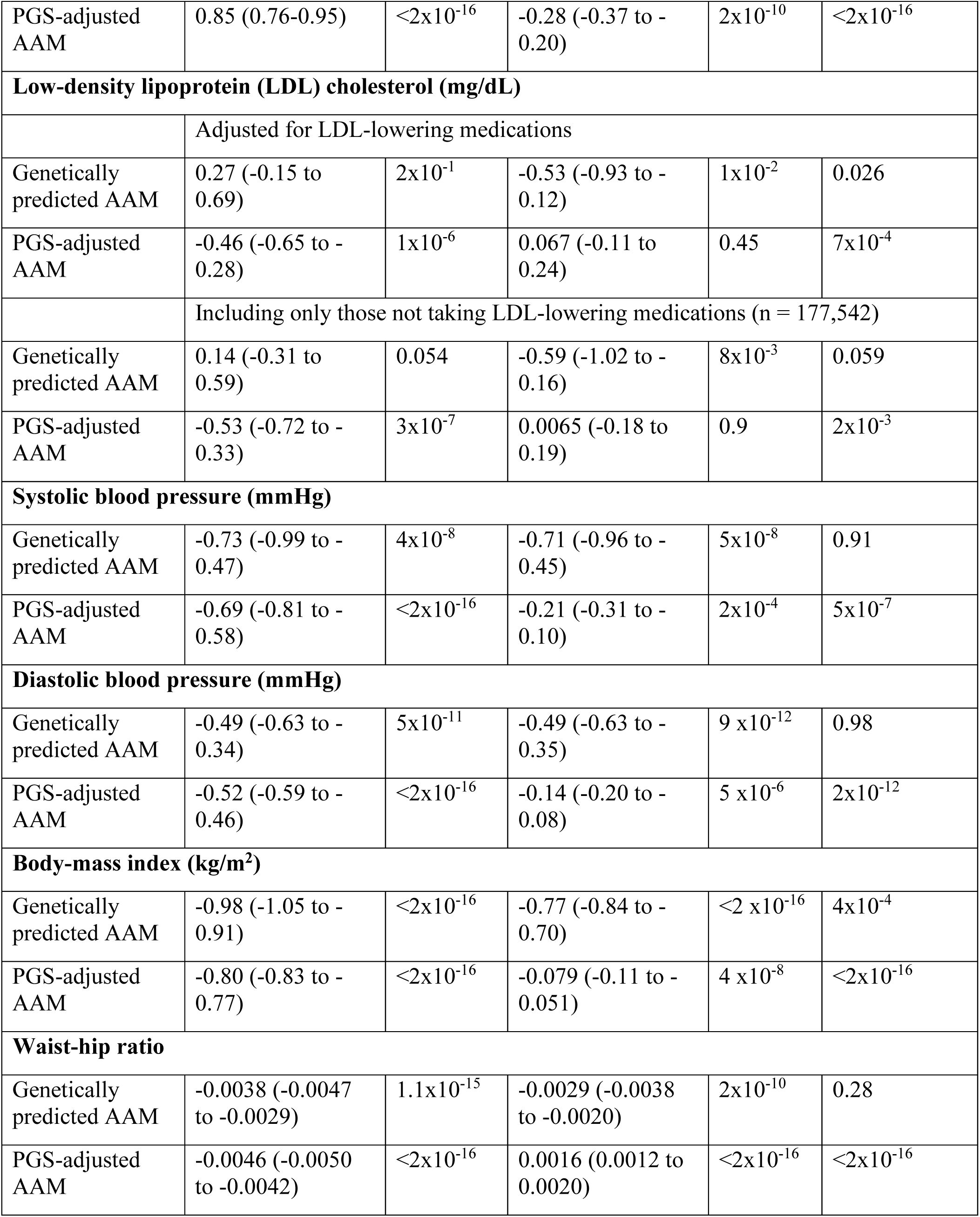
Associations of genetically predicted and PGS-adjusted AAM with coronary artery disease and its risk factors. AAM: age at menarche; PGS: polygenic score

| Coronary artery disease |  |  |  |  |  |
| --- | --- | --- | --- | --- | --- |
|  | Change per year increase in AAM |  |  |  |  |
|  | Values earlier than the mean |  | Values later than the mean |  | *p* for difference between slopes |
|  | Odds ratio (95% CI) | *p* | Odds ratio (95% CI) | *p* |  |
| Genetically predicted AAM | 0.91 (0.83 to 0.99) | $3 \times 10^{-2}$ | 0.92 (0.84 to 1) | 0.063 | 0.87 |
| PGS-adjusted AAM | 0.92 (0.88 to 0.95) | $3 \times 10^{-6}$ | 1.11 (1.08 to 1.15) | $2 \times 10^{-9}$ | $7 \times 10^{-10}$ |
| Coronary artery disease risk factors |  |  |  |  |  |
|  | Change per year increase in AAM |  |  |  |  |
|  | Values earlier than the mean |  | Values later than the mean |  | *p* for difference between slopes |
|  | Slope (95% CI) | *p* | Slope (95% CI) | *p* |  |
| Hemoglobin A1c (%) |  |  |  |  |  |
| Genetically predicted AAM | -0.034 (-0.041 to -0.027) | $< 2 \times 10^{-16}$ | -0.01 (-0.02 to -0.006) | $3 \times 10^{-3}$ | $6 \times 10^{-5}$ |
| PGS-adjusted AAM | -0.022 (-0.025 to -0.019) | $< 2 \times 10^{-16}$ | 0.007 (0.004 to 0.01) | $1.3 \times 10^{-6}$ | $< 2 \times 10^{-16}$ |
| Triglycerides (mg/dL) |  |  |  |  |  |
| Genetically predicted AAM | -2.82 (-3.84 to -1.79) | $7 \times 10^{-8}$ | -2.63 (-3.62 to -1.64) | $2 \times 10^{-7}$ | 0.83 |
| PGS-adjusted AAM | -4.25 (-4.7 to -3.8) | $< 2 \times 10^{-16}$ | 1.2 (0.78 to 1.62) | $2 \times 10^{-8}$ | $< 2 \times 10^{-16}$ |
| High-density lipoprotein (HDL) cholesterol (mg/dL) |  |  |  |  |  |
| Genetically predicted AAM | 0.90 (0.69-1.11) | $< 2 \times 10^{-16}$ | 0.74 (0.54-0.95) | $8 \times 10^{-13}$ | 0.38 |
| PGS-adjusted AAM | 0.85 (0.76-0.95) | $<2 \times 10^{-16}$ | -0.28 (-0.37 to -0.20) | $2 \times 10^{-10}$ | $<2 \times 10^{-16}$ |
| **Low-density lipoprotein (LDL) cholesterol (mg/dL)** |  |  |  |  |  |
|  | Adjusted for LDL-lowering medications |  |  |  |  |
| Genetically predicted AAM | 0.27 (-0.15 to 0.69) | $2 \times 10^{-1}$ | -0.53 (-0.93 to -0.12) | $1 \times 10^{-2}$ | 0.026 |
| PGS-adjusted AAM | -0.46 (-0.65 to -0.28) | $1 \times 10^{-6}$ | 0.067 (-0.11 to 0.24) | 0.45 | $7 \times 10^{-4}$ |
|  | Including only those not taking LDL-lowering medications (n = 177,542) |  |  |  |  |
| Genetically predicted AAM | 0.14 (-0.31 to 0.59) | 0.054 | -0.59 (-1.02 to -0.16) | $8 \times 10^{-3}$ | 0.059 |
| PGS-adjusted AAM | -0.53 (-0.72 to -0.33) | $3 \times 10^{-7}$ | 0.0065 (-0.18 to 0.19) | 0.9 | $2 \times 10^{-3}$ |
| **Systolic blood pressure (mmHg)** |  |  |  |  |  |
| Genetically predicted AAM | -0.73 (-0.99 to -0.47) | $4 \times 10^{-8}$ | -0.71 (-0.96 to -0.45) | $5 \times 10^{-8}$ | 0.91 |
| PGS-adjusted AAM | -0.69 (-0.81 to -0.58) | $<2 \times 10^{-16}$ | -0.21 (-0.31 to -0.10) | $2 \times 10^{-4}$ | $5 \times 10^{-7}$ |
| **Diastolic blood pressure (mmHg)** |  |  |  |  |  |
| Genetically predicted AAM | -0.49 (-0.63 to -0.34) | $5 \times 10^{-11}$ | -0.49 (-0.63 to -0.35) | $9 \times 10^{-12}$ | 0.98 |
| PGS-adjusted AAM | -0.52 (-0.59 to -0.46) | $<2 \times 10^{-16}$ | -0.14 (-0.20 to -0.08) | $5 \times 10^{-6}$ | $2 \times 10^{-12}$ |
| **Body-mass index (kg/m<sup>2</sup>)** |  |  |  |  |  |
| Genetically predicted AAM | -0.98 (-1.05 to -0.91) | $<2 \times 10^{-16}$ | -0.77 (-0.84 to -0.70) | $<2 \times 10^{-16}$ | $4 \times 10^{-4}$ |
| PGS-adjusted AAM | -0.80 (-0.83 to -0.77) | $<2 \times 10^{-16}$ | -0.079 (-0.11 to -0.051) | $4 \times 10^{-8}$ | $<2 \times 10^{-16}$ |
| **Waist-hip ratio** |  |  |  |  |  |
| Genetically predicted AAM | -0.0038 (-0.0047 to -0.0029) | $1.1 \times 10^{-15}$ | -0.0029 (-0.0038 to -0.0020) | $2 \times 10^{-10}$ | 0.28 |
| PGS-adjusted AAM | -0.0046 (-0.0050 to -0.0042) | $<2 \times 10^{-16}$ | 0.0016 (0.0012 to 0.0020) | $<2 \times 10^{-16}$ | $<2 \times 10^{-16}$ |

In contrast, the association between PGS-adjusted AAM and risk of CAD was U-shaped, with both earlier and later values associated with increased CAD risk (Figure 2). For PGS-adjusted AAM that was earlier than the mean, each 1-year increase (causing AAM to be less early) was associated with an OR for CAD of 0.91 (Table 1). In other words, in women whose PGS-adjusted AAM was earlier than the mean, for every 1 year that AAM was less early due to factors other than the PGS, the odds of CAD were lower by 9%. In contrast, for PGS-adjusted AAM later than the mean, each 1-year increase (causing AAM to be even later) was associated with an OR for CAD of 1.11, i.e., the odds of CAD were higher by 11% (Table 1).

Because the 2017 AAM GWAS meta-analysis included data from the UK Biobank, which could introduce bias in the above analyses, we conducted sensitivity analyses using the results from an AAM GWAS meta-analysis published in 2014 that did not include the UK Biobank. These analyses showed similar results for CAD (Supplemental Figure 3, Supplemental Table 1, (29)). We also conducted sensitivity analyses excluding women with extreme values of AAM (at the upper and lower 0.1%) and obtained similar results.

### Hemoglobin A1c

We then examined the associations of genetically predicted AAM and PGS-adjusted AAM with CAD risk factors. Just as the relationship between genetically predicted AAM and CAD was linear, we observed linear (or roughly linear) relationships between genetically predicted AAM and most CAD risk factors. In contrast, for PGS-adjusted AAM we observed mostly non-linear relationships with CAD risk factors.

For hemoglobin A1c, an indicator of average blood glucose over the preceding 3 months, genetically predicted AAM demonstrated a roughly negative linear relationship, with later genetically predicted AAM associated with lower HbA1c (Figure 3). Linear spline analyses indicated that while both slopes were negative, for values of genetically predicted AAM earlier than the mean, the slope of the association with HbA1c was slightly steeper than for values of genetically predicted AAM later than the mean (-0.034 and -0.022 %/year, respectively; *p* for difference between slopes: 7 x 10^-5^; Table 1). In contrast, PGS-adjusted AAM demonstrated a U-shaped relationship with HbA1c: both earlier and later PGS-adjusted AAM were associated with an increase in HbA1c (for values earlier than the mean: slope = -0.022 %/year, for values later than the mean: slope = 0.007%/year; *p* for difference between slopes < 2 x 10^-16^; Figure 3, Table 1). This was similar to the U-shaped relationship of PGS-adjusted AAM with CAD risk.

**Figure 3:**
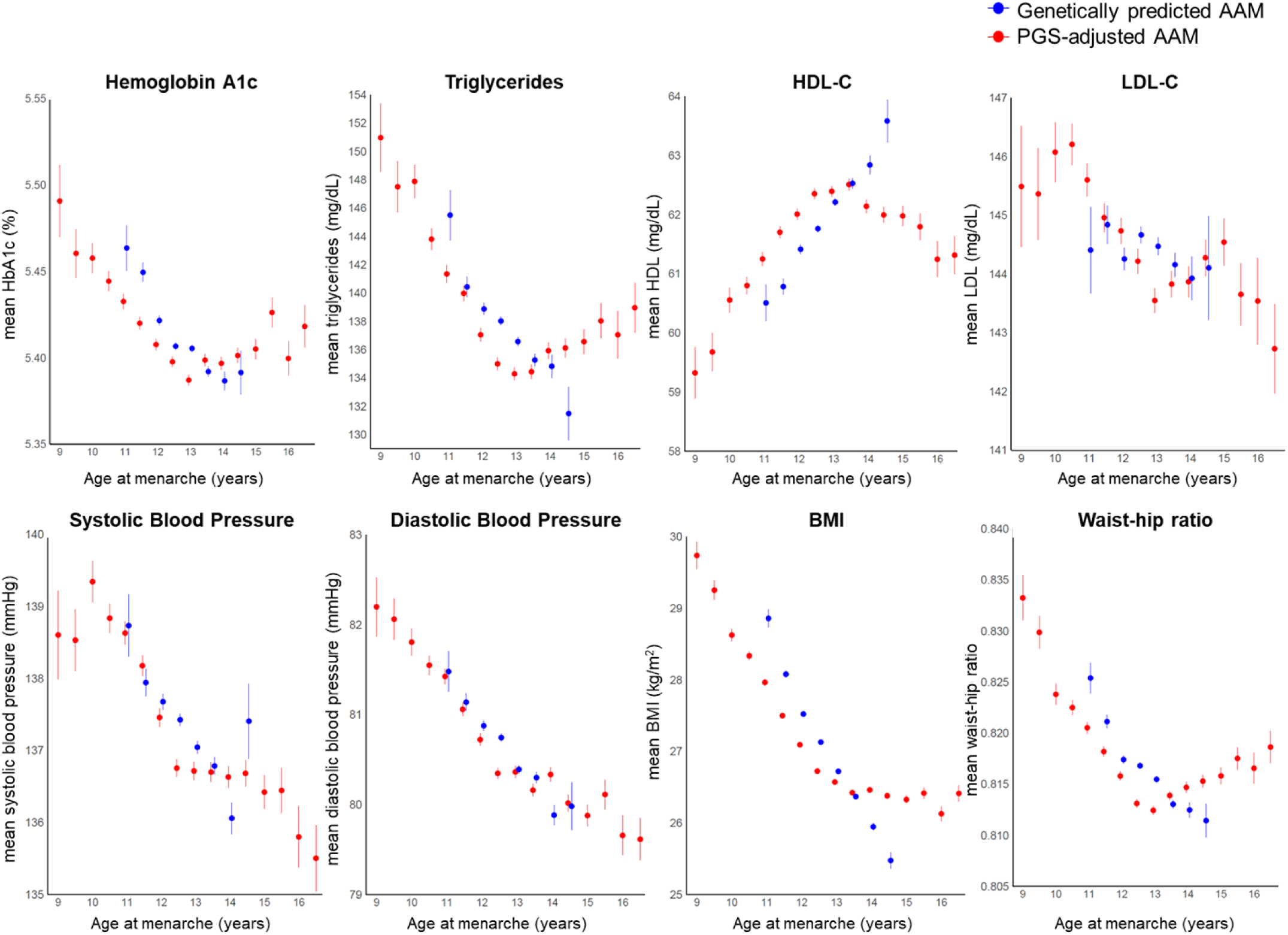
Associations of genetically predicted and PGS-adjusted AAM with risk factors for CAD in the UK Biobank. We observed linear (or roughly linear) relationships between genetically predicted AAM and most CAD risk factors. In contrast, for PGS-adjusted AAM we observed mostly non-linear relationships with CAD risk factors. The U-shaped relationship of PGS-adjusted AAM with CAD was mirrored by the relationships with HbA1c, triglycerides, HDL-C, and waist-hip ratio. This raises the possibility that dysglycemia, dyslipidemia and central adiposity may mediate the relationship between later AAM and increased risk of CAD. For LDL-C, n = 177,542 after excluding those taking LDL-C lowering medications. Dots represent estimates, bars represent standard errors. AAM: age at menarche; BMI: body mass index; CAD: coronary artery disease; HDL-C: high-density lipoprotein cholesterol; LDL-C: low-density lipoprotein cholesterol; PGS: polygenic score

Comparing genetically predicted AAM and PGS-adjusted AAM, for values earlier than the mean, the slopes for the associations with HbA1c were comparable (Table 1). However, for values of AAM later than the mean, different slopes were seen for genetically predicted vs. PGS-adjusted AAM; as noted above, for every 1 year that AAM was delayed, HbA1c *decreased* by 0.01% for genetically predicted AAM but *increased* by 0.007% for PGS-adjusted AAM.

### Lipids

Genetically predicted AAM demonstrated linear relationships with triglycerides and HDL-C (Figure 3), with later genetically predicted AAM associated with lower triglycerides and higher HDL-C (Figure 3) and no significant difference in slopes between earlier and later genetically predicted AAM (*p* for difference in slopes for triglycerides = 0.83 mg/dL/year, for HDL-C = 0.38 mg/dL/year; Table 1). In contrast, PGS-adjusted AAM demonstrated non-linear relationships with HDL-C and triglycerides – U-shaped for triglycerides and inverted-U-shaped for HDL-C (Figure 3) - with both increasingly earlier and later PGS-adjusted AAM associated with higher triglycerides and lower HDL-C (Figure 3, Table 1).

For LDL-C, associations showed a less clear pattern. Earlier genetically predicted AAM showed no association with LDL-C, and later genetically predicted AAM showed a negative linear relationship (Table 1). In contrast, for PGS-adjusted AAM, earlier values showed a negative linear relationship, but later PGS-adjusted AAM showed no significant association (Figure 3, Table 1). Similar results were seen when analyses excluded those taking LDL-lowering medications (Table 1).

### Blood pressure

Later genetically predicted AAM was associated with both lower SBP and lower DBP (Figure 3), with no significant difference between the slopes for values of genetically predicted AAM earlier vs. later than the mean (Table 1). Later PGS-adjusted AAM was also associated with lower SBP and DBP (Figure 3), but the slopes of the associations were steeper for values of PGS-adjusted AAM that were earlier vs. later than the mean (Table 1).

### Adiposity

We studied two estimates of adiposity: BMI and waist-hip ratio. For BMI, we found a roughly negative linear relationship between genetically predicted AAM and BMI, with later genetically predicted AAM associated with lower BMI (Figure 3, Table 1). The slope of the association with BMI was slightly steeper for values of genetically predicted AAM earlier vs. later than the mean (Table 1). This pattern was similar to the relationship seen between genetically predicted AAM and HbA1c.

For PGS-adjusted AAM, a different relationship was seen with BMI (Figure 3). While later PGS-adjusted AAM was consistently associated with lower BMI, the slope of the association was ten-fold steeper for values earlier vs. later than the mean (-0.80 kg/m2/year vs. -0.079 kg/m2/year respectively; *p* for difference between slopes 2 x 10^-16^; Table 1). This resembled the patterns seen with SBP and DBP.

For waist-hip ratio, genetically predicted AAM showed a negative linear association, with no significant difference in slopes for the association when genetically predicted AAM was earlier vs. later than the mean (Figure 3, Table 1). In contrast, the relationship of PGS-adjusted AAM with waist-hip ratio was U-shaped, with values both earlier and later than the mean associated with higher waist-hip ratio (Figure 3, Table 1), similar to the associations seen with CAD, HbA1c, triglycerides, and HDL-C.

### Validation in the Women’s Genome Health Study

Validation studies using data from the WGHS produced results similar to the above results for the UK Biobank. In the WGHS, genetically predicted AAM showed linear relationships with HbA1c, triglycerides, HDL-C, SBP, DBP, BMI and waist-hip ratio and no clear relationship with LDL-C (Figure 4). Just as in the UK Biobank, PGS-adjusted AAM showed U-shaped relationships with triglycerides and HbA1c, an inverted-U-shaped relationship with HDL-C, and a not completely linear relationship with BMI in the WGHS (Figure 4).

**Figure 4:**
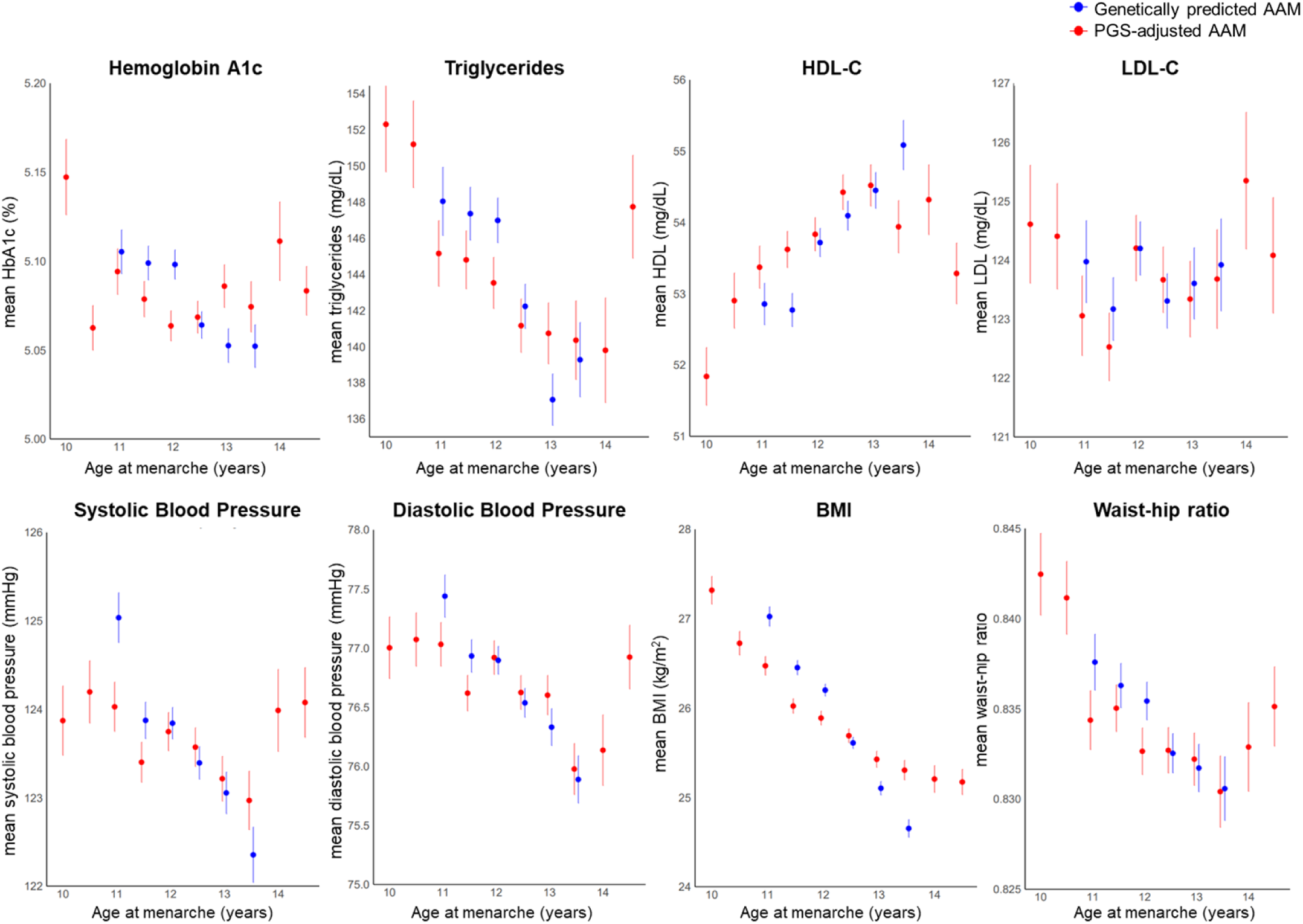
Associations of genetically predicted and PGS-adjusted AAM with risk factors for CAD in the WGHS. genetically predicted AAM showed linear or roughly linear associations with most CAD risk factors and PGS-adjusted AAM showed mostly non-linear associations with most CAD risk factors. Similar to results in the UK Biobank, hemoglobin A1c, triglycerides and HDL-C showed U-shaped associations with PGS-adjusted AAM which support conclusions from the primary analyses that dysglycemia, dyslipidemia and central adiposity may mediate the relationship between later AAM and increased risk of CAD. For LDL-C, n = 22,495 after excluding those taking cholesterol-lowering medications. Dots represent estimates, bars represent standard errors. AAM: age at menarche; BMI: body mass index; CAD: coronary artery disease; HDL-C: high density lipoprotein cholesterol; LDL-C: low density lipoprotein cholesterol; PGS: polygenic score; WGHS: Women’s Genome Health Study.

There were two differences between results from the WGHS and the UK Biobank: 1) for SBP and DBP, PGS-adjusted AAM showed roughly U-shaped relationships in the WGHS (Figure 4) rather than the not fully linear relationships seen in the UK Biobank (Figure 3), and 2) for waist-hip ratio, PGS-adjusted AAM showed a negative linear relationship for values earlier than the mean but no significant relationship for values later than the mean in the WGHS (Figure 4) compared to the clear U-shaped relationship seen in the UK Biobank (Figure 2).

For risk of CAD itself, the relationship of genetically predicted AAM with CAD in the WGHS was similar to that in the UK Biobank (Figure 5, Supplemental Table 1, (29)), with a negative linear relationship in both cohorts. For the relationship of PGS-adjusted AAM with CAD, analyses in the WGHS demonstrated a reverse-J shaped relationship (Figure 5, Supplemental Table 1, (29)), slightly different from the U-shaped relationship seen in the UK Biobank.

**Figure 5:**
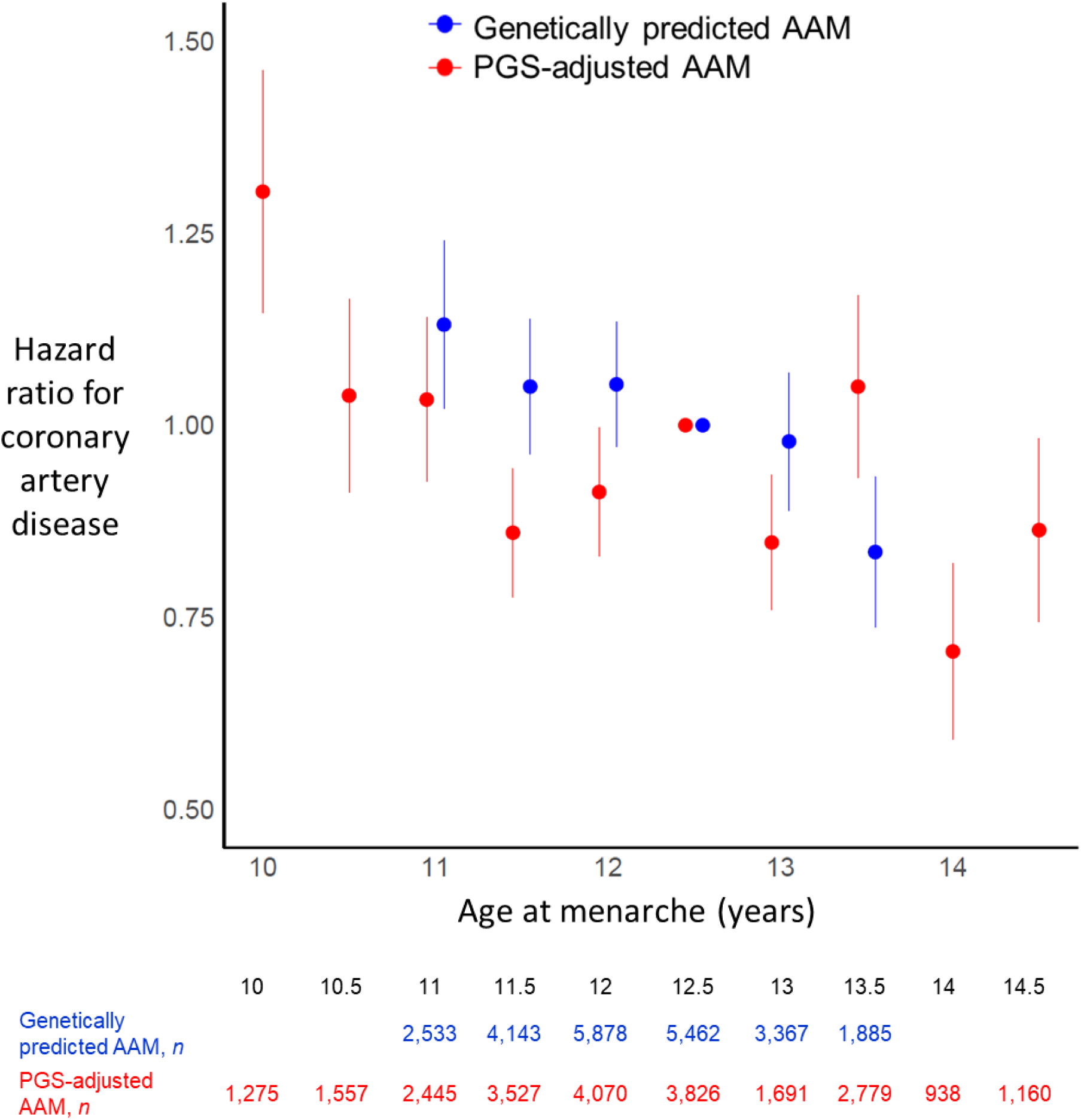
Associations of genetically predicted and PGS-adjusted AAM with hazard ratio for risk of CAD in the WGHS. Genetically predicted AAM demonstrated a negative linear association with risk of CAD, similar to the results in the UKBB. PGS-adjusted AAM demonstrated a reverse-J shaped relationship, slightly different from the U-shaped relationship seen in the UK Biobank. Analyses in the WGHS largely validated the results from the UK Biobank. The different associations of later PGS-adjusted AAM with risk of CAD may have been due to differences in environmental factors between the two study cohorts. AAM: age at menarche; ΔAAM: change in AAM; CAD: coronary artery disease; PGS: polygenic score. Dots represent estimates, bars represent standard errors.

## DISCUSSION

In this study, we dissected variation in AAM into variation attributable to common genetic variants, as estimated by a PGS, (as reflected by genetically predicted AAM), and variation adjusted for the PGS (as reflected by PGS-adjusted AAM), and we found different relationships with these two sources of variation in AAM with risk of CAD and with CAD risk factors, particularly when causing AAM to occur later.

In general, later AAM showed favorable associations when attributable to common genetic variation (as estimated by the PGS) and harmful or neutral associations when attributable to other yet-to-be-identified sources of variation. If all sources of variation were affecting CAD and CAD risk factors wholly through AAM itself, the associations would be similar regardless of the source of variation studied. Our finding that these associations varied based on the underlying source of variation driving later AAM therefore indicates that it is not later AAM itself that causes increased risk of CAD. Rather, there appear to be PGS-independent factors that cause both later AAM and increased risk of CAD and unfavorable cardiometabolic risk profiles. Such factors could include environmental or acquired influences (such as chronic illnesses, chronic stress, undernutrition) as well as genetic influences not captured by the PGS, and future studies will identify these factors and determine how they contribute to CAD risk.

An alternative possibility is that later AAM itself does in fact cause increased risk of CAD, regardless of underlying source of variation. For this to be true, given our finding that later genetically predicted AAM is associated with lower CAD risk, there would have to be strong direct (pleiotropic) effects of common genetic variants on risk of CAD, with later genetically predicted AAM strongly protective against CAD, such that the net effect is the lower risk of CAD that we observed with later genetically predicted AAM. However, previous MR studies have suggested that such pleiotropic effects of common genetic variants for AAM on risk of CAD are small (10,31–33). Hence, this alternative possibility is not supported by existing evidence, and the more likely explanation is that PGS-independent factors that cause later AAM have direct deleterious effects on CAD risk factors and CAD.

Earlier AAM, whether attributable to common genetic variants or other sources of variation, was consistently associated with greater cardiometabolic risk. Thus, it seems that earlier AAM itself is intrinsically associated with risk for CAD and worsening CAD risk factors; indeed, some Mendelian randomization studies suggest that earlier AAM is causative of these negative outcomes (11,31). However, other Mendelian randomization studies have suggested that, because many loci that affect AAM also affect BMI, the associations with increased risk of CAD are mainly through effects on BMI, and AAM itself may have only a small direct influence (10,33).

The U-shaped relationship of PGS-adjusted AAM with CAD was mirrored by the relationships with HbA1c, triglycerides, HDL-C, and waist-hip ratio. This raises the possibility that dysglycemia, dyslipidemia, and central adiposity contribute to the relationship between later AAM and increased risk of CAD; further studies will be required to formally evaluate these CAD risk factors as potential mediators of this relationship. As noted above, previous studies evaluating associations of later AAM with these CAD risk factors have had conflicting results.

While differences between study cohorts may have accounted for some of the differing results, our findings raise the additional possibility that the relationships between later AAM and these outcomes may have been obscured by opposing effects of genetically predicted and PGS-adjusted variation in AAM. Of note, the associations with waist-hip ratio differed from those with BMI, suggesting that central adiposity, reflected by waist-hip ratio, is more relevant than BMI for CAD, as has been suggested by prior studies (34,35).

Given the well-known effects of increased adiposity on CAD and CAD risk factors such as dysglycemia and dyslipidemia, it is possible that associations of PGS-adjusted variation in AAM with CAD, dysglycemia, and dyslipidemia are largely mediated by associations with BMI and central adiposity; however, we were unable to test this possibility with UK Biobank data. The relationship between AAM and BMI is complex because childhood BMI – not available in the UK Biobank – affects both AAM and adult BMI, and AAM is itself also associated with adult BMI. Thus, adjusting for adult BMI would introduce collider bias. Future work could focus on disentangling these associations by using cohorts with measures of childhood BMI and/or genetic tools such as clustering analyses to generate partitioned polygenic scores (36,37).

Our analyses in the Women’s Genome Health Study (WGHS) largely validated our results from the UK Biobank. Genetically predicted AAM demonstrated linear relationships with CAD and CAD risk factors, and PGS-adjusted AAM demonstrated mostly non-linear relationships with these outcomes, supporting the conclusions described earlier. However, the two cohorts also demonstrated some differences, most notably in the association of PGS-adjusted AAM with the risk of CAD and waist-hip ratio. In the UK Biobank, PGS-adjusted *later* AAM was associated with an increased risk of CAD, but there was no association in the WGHS. For waist-hip ratio, PGS-adjusted AAM demonstrated a clear U-shaped association in the UK Biobank but a reverse J-shaped relationship in the WGHS. There are several potential reasons for this difference. First, it is possible that the smaller sample size and lower power in the WGHS affected the ability to find significant associations with PGS-adjusted AAM and risk of CAD and waist-hip ratio.

Second, environmental influences affecting AAM (which would contribute to PGS-adjusted AAM) could differ between the two cohorts. The WGHS recruited women in the United States of America born in 1950 or earlier, while the UK Biobank recruited women in the UK who were born between 1932 and 1969. The different impact of global events such as World War II on the two countries could contribute to differences in the PGS-independent factors (which includes environmental factors) and, in turn, to different associations with risk of CAD. Third, the WGHS excluded women with a history of CAD at the time of enrollment whereas the UK Biobank did not, and this may also have led to differences in PGS-adjusted factors between the cohorts.

Fourth, the participants in the WGHS were health professionals while the UK Biobank drew from the general UK population, and this may have led to further differences in PGS-adjusted factors, such as higher socioeconomic status, greater knowledge of CAD and its risk factors, healthier diets and lifestyles, and use of preventative interventions, as well as potentially less variation in these factors. While the analyses also differed in the CAD measure used from each cohort – prevalence of CAD was analyzed in the UK Biobank compared to incidence of CAD in the WGHS – it is unlikely to account for the difference in results, as an analysis of incident CAD in the Million Women Study in the UK also showed a U-shaped relationship between AAM and CAD (1). Despite these differences in the results between the two cohorts, results from both cohorts consistently demonstrated differences between the associations of *earlier* vs. *later* PGS-adjusted AAM with these outcomes.

In 2021, Liang et al. used a PGS to represent genetically predicted AAM and examined associations with all-cause mortality, also using data from the UK Biobank. Interestingly, they found a U-shaped association with mortality, with both earlier and later genetically predicted AAM associated with higher risk; this finding contrasts with the linear relationship we found between genetically predicted AAM and CAD. This difference suggests that later genetically predicted AAM increases the risk of causes of mortality other than CAD, and future studies of genetically predicted AAM are needed to identify these causes.

Prior studies that have also dissected influences on human traits into genetic vs. environmental influences have found differences in the magnitude of effect of genetics vs. environmental influences on health outcomes (21,22). Interestingly, our results show not only different magnitudes of association of genetically predicted and PGS-adjusted AAM with risk of CAD, but also opposite directionality. This further underscores the value of separating the effects of genetics vs. other influences while studying human traits as they can have starkly different effects.

Sensitivity analyses using a GWAS that did not include the primary cohort, the UK Biobank, produced similar results and suggested that these results were not biased by overfitting.

Additionally, sensitivity analyses excluding women with extreme values of AAM also produced similar results, suggesting that results were not heavily influenced by outliers.

One limitation of this study is that while the PGS represents genetic influences on AAM, it does not represent all genetic factors that influence AAM. Our analyses demonstrated that the PGS explains 15.8% of the variation in the observed AAM, but prior studies suggest that 49-73% of variation in AAM is inherited (18,19); hence, a large amount of the variation in AAM due to genetic factors remains unexplained. Another limitation of this study is that the PGS represents just one method of capturing the effects of common genetic variants that affect AAM. There may be several pathways causing later AAM represented within these common genetic variants, and using a single PGS to represent all those effects may obscure relationships with each individual pathway. Future studies may identify these different pathways by methods such as clustering analyses (36,37), which would then allow an estimation of multiple polygenic scores, each representing a different pathway, to study their associations with risk of CAD and CAD risk factors.

Distinguishing between sources of variation in AAM has provided a novel lens through which to study associations of AAM with CAD and has allowed us to uncover differences in the associations of genetically predicted vs. PGS-adjusted AAM with CAD and CAD risk factors.

Because later puberty in women, but not men, has been associated with an increased risk of CAD, studying these differences further may provide unique insights into mechanisms that affect CAD risk specifically in women.

## Data Availability

The data that support the findings of this study are available through the UK Biobank and Women’s Genome Health Study.

## ACKNOWLEDGEMENTS

The authors thank Evan Schafer and Jia Zhu for their support with statistical analyses. This research has been conducted using data from UK Biobank, a major biomedical database: www.ukbiobank.ac.uk

## SOURCES OF FUNDING

A.S. was supported by National Institutes of Health (NIH) grant T32DK007699. The WGHS and its parent cohort, the Women’s Health Study (WHS), have been supported by the National Heart, Lung, and Blood Institute (HL043851 and HL080467) and the National Cancer Institute (CA047988 and UM1CA182913), with support and funding for genotyping provided by Amgen. Some cardiovascular endpoints in the WHS were funded by HL099355.

## DISCLOSURE SUMMARY

A.S. has no relevant disclosures or relationships with industry. D.I.C has no relevant disclosures or relationships with industry. Y.-M.C. receives royalties from UpToDate on topics related to puberty.

## Non-standard Abbreviations and Acronyms

AAM: age at menarche
BMI: body-mass index
CAD: coronary artery disease
DBP: diastolic blood pressure
HbA1c: hemoglobin A1c
HDL-C: high-density lipoprotein cholesterol
LDL-C: low-density lipoprotein cholesterol
PGS: polygenic score
SBP: systolic blood pressure
WGHS: Women’s Genome Health Study

## Notes

### Funding Statement

A.S. was supported by National Institutes of Health (NIH) grant T32DK007699. The Women’s Genome Health Study and its parent cohort, the Women’s Health Study (WHS), have been supported by the National Heart, Lung, and Blood Institute (HL043851 and HL080467) and the National Cancer Institute (CA047988 and UM1CA182913), with support and funding for genotyping provided by Amgen. Some cardiovascular endpoints in the WHS were funded by HL099355.

### Author Declarations

Boston Children’s Hospital Institutional Review Board

### Summary of Updates

The variables and terminology used to describe the predictor variables in the manuscript text have been updated to improve clarity. Specifically, the terminology has been updated to genetically predicted AAM and PGS-adjusted AAM. The figures have been updated to reflect this. The data and core analyses remain the same. Discussion has been updated to include limitations of adjusting for BMI.

