## supplemental file for "Age at Menarche and Coronary Artery Disease Risk: Divergent Associations with Different Sources of Variation"

**SUPPLEMENTAL MATERIAL**

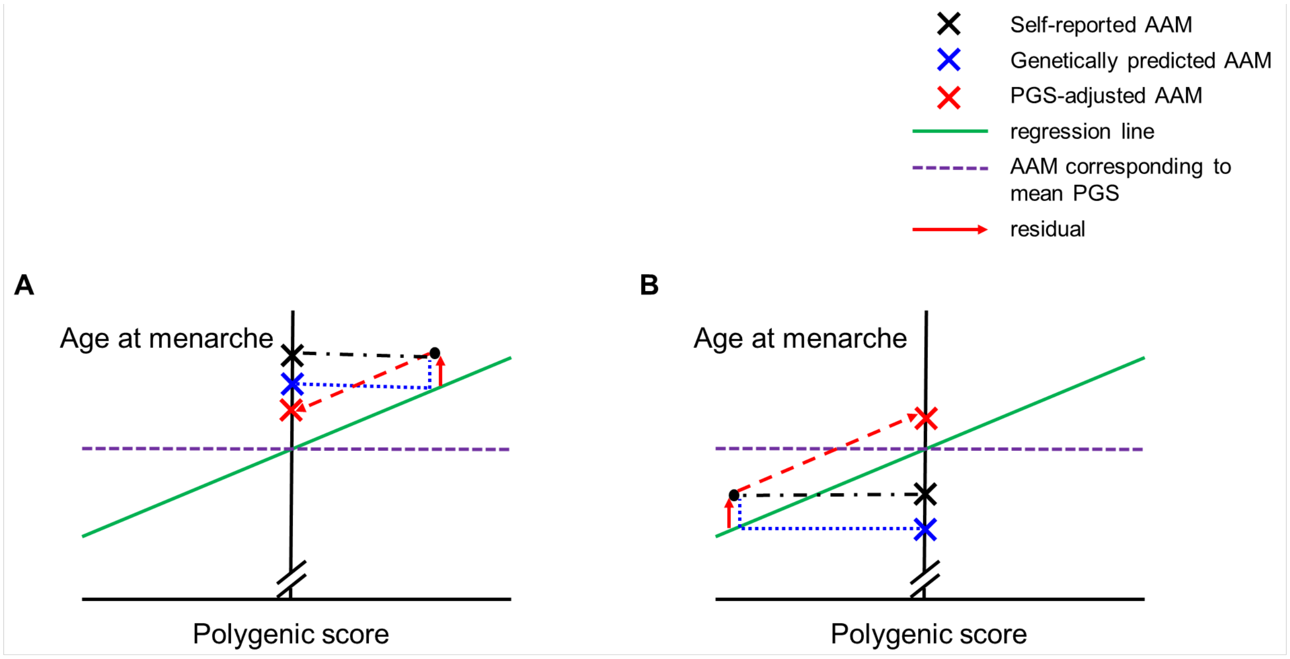


**Supplemental Figure 1: Derivation of genetically predicted and PGS-adjusted age at menarche.** For all women in the dataset, self-reported age at menarche (AAM) was regressed against a polygenic score (PGS) for AAM. The green line indicates the regression line for self-reported AAM against PGS, the horizontal dashed purple line represents the observed AAM at the mean PGS for the cohort (which is 12.95 years in the UK Biobank cohort). Panels A and B represent examples of two different women in the cohort. In each panel, the black dot represents an example of a single woman; the black X and the black dashed line indicate the woman's self-reported AAM; the blue X indicates the woman's genetically predicted AAM, with the blue dashed lines representing its derivation based on the regression; the red X indicates the woman's PGS-adjusted AAM, calculated by adding the residual (red solid arrow) to the AAM at the mean PGS for the cohort, with the red dashed arrow representing its derivation. For example, in panel A, the self-reported AAM is 16 years, the genetically predicted AAM is 13.42 years, and the

PGS-adjusted AAM is 15.53 years; in panel B, the self-reported AAM is 12 years, the genetically predicted AAM is 10.99 years, and the PGS-adjusted AAM is 13.94 years.

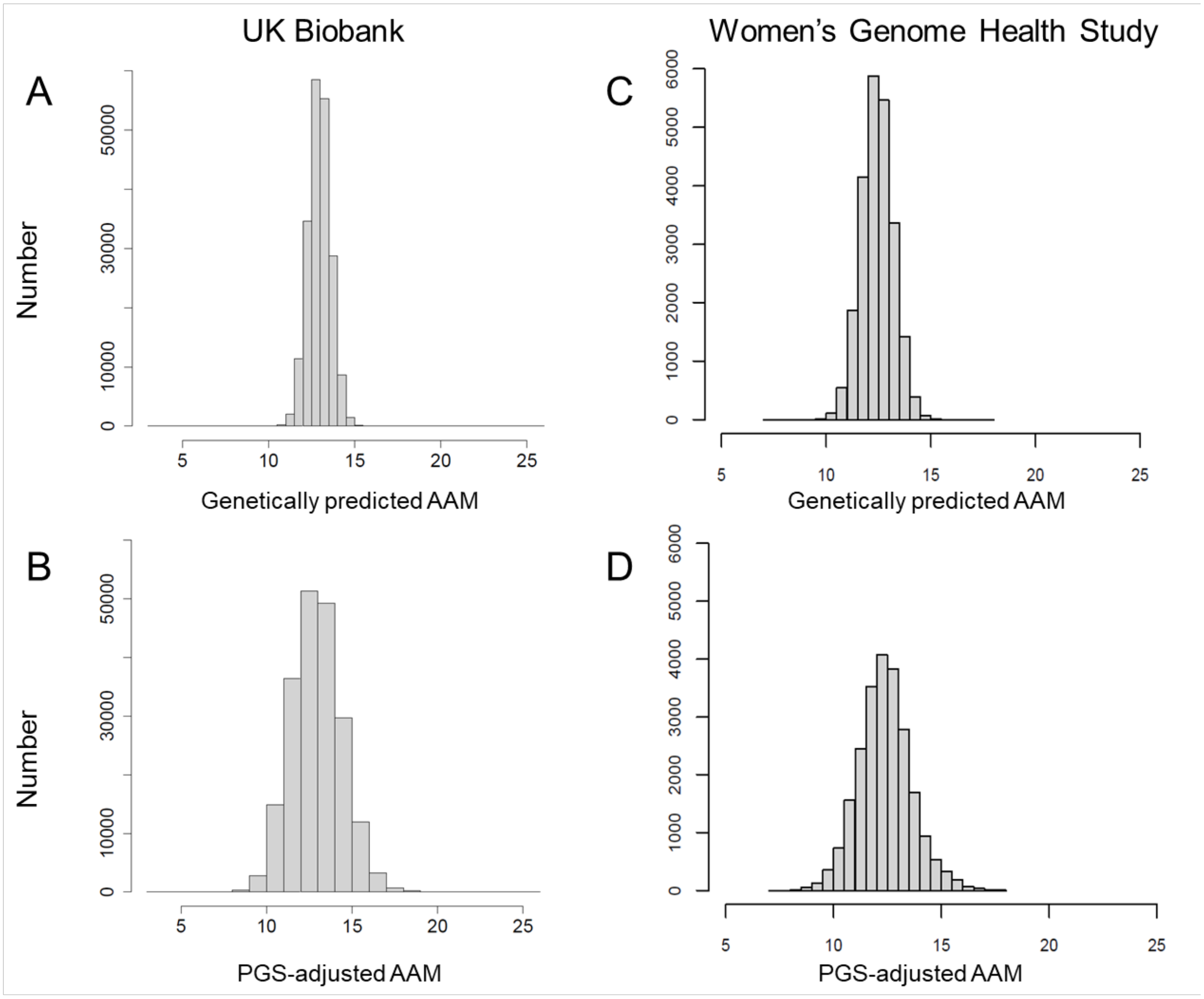


**Supplemental Figure 2: Distribution of genetically predicted AAM and PGS-adjusted AAM in study cohorts.** Panels show genetically predicted AAM in the UK Biobank (A), PGS-adjusted AAM in the UK Biobank (B), genetically predicted AAM in the WGHS (C), and PGS-adjusted AAM in the WGHS (D).

AAM: age at menarche; PGS: polygenic score, WGHS: Women's Genome Health Study.

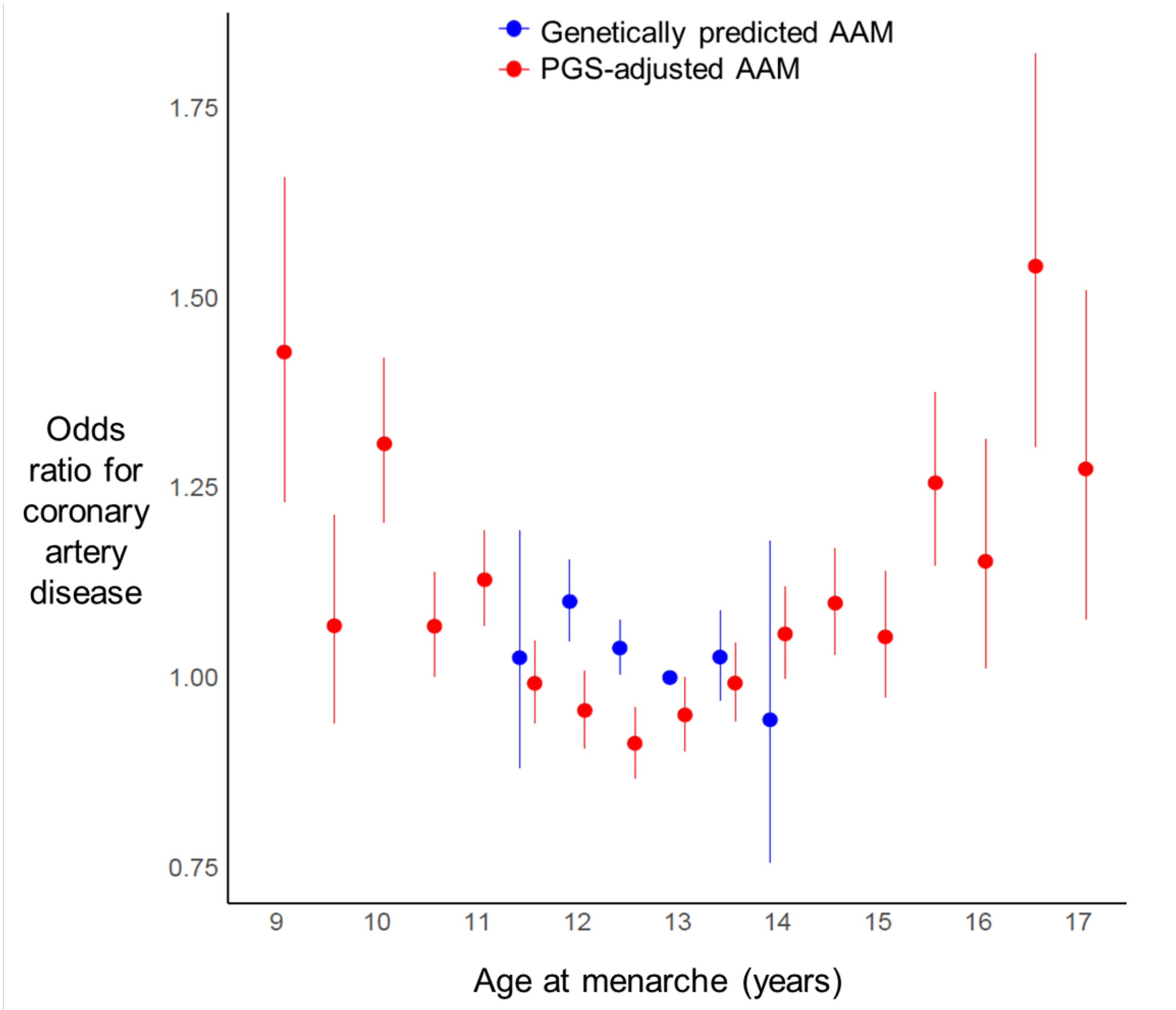


| | 9 | 9.5 | 10 | 10.5 | 11 | 11.5 | 12 | 12.5 | 13 | 13.5 | 14 | 14.5 | 15 | 15.5 | 16 | 16.5 | 17 |
|---|---|---|---|---|---|---|---|---|---|---|---|---|---|---|---|---|---|
| Genetically predicted AAM, *n* | | | | | | 2,024 | 24,913 | 83,074 | 73,186 | 16,838 | 1,002 | | | | | | |
| PGS-adjusted AAM, *n* | 1,531 | 2,750 | 5,733 | 12,656 | 17,159 | 19,728 | 22,096 | 25,781 | 24,221 | 22,130 | 17,048 | 12,397 | 7,976 | 4,913 | 2,474 | 1,126 | 1,318 |

**Supplemental Figure 3:** Association of genetically predicted and PGS-adjusted age at menarche with coronary artery disease in the UK Biobank using a PGS based on a 2014 genome-wide association study meta-analysis that did not include data from the UK Biobank.

AAM: age at menarche; PGS: polygenic score. Dots represent estimates, bars represent standard errors.

| | **Change per year increase in AAM** | | | | |
|---|---|---|---|---|---|
| | **Values earlier than the mean** | | **Values later than the mean** | | ***p* for difference between slopes** |
| **Coronary artery disease** | | | | | |
| | Odds ratio (95% CI) | *p* | Odds ratio (95% CI) | *p* | |
| Genetically predicted AAM | 0.89 (0.77 to 1.04) | 0.14 | 0.98 (0.86 to 1.1) | 0.79 | 0.43 |
| PGS-adjusted AAM | 0.91 (0.88 to 0.94) | $5x10^{-7}$ | 1.09 (1.06 to 1.13) | $4x10^{-7}$ | $5x10^{-9}$ |

**Supplemental Table 1:** Associations between genetically predicted and PGS-adjusted age at menarche and coronary artery disease using a PGS based on a 2014 genome-wide association study meta-analysis that did not include data from the UK Biobank.

AAM: age at menarche; PGS: polygenic score
